# Improving postpartum hemorrhage risk prediction using longitudinal electronic medical records

**DOI:** 10.1101/2021.03.16.21253746

**Authors:** Amanda B Zheutlin, Luciana Vieira, Shilong Li, Zichen Wang, Emilio Schadt, Susan Gross, Joanne Stone, Eric Schadt, Li Li

## Abstract

**Objective:** Postpartum hemorrhage (PPH) remains a leading cause of preventable maternal mortality in the US. Our goal was to develop a novel risk assessment tool and compare its accuracy to those used in current practice.

**Materials and Methods:** We used a PPH digital phenotype we developed and validated previously to identify 6,639 cases from our delivery cohort (N=70,948). Using a vast array of known and potential risk factors extracted from electronic medical records available prior to delivery, we trained a gradient boosting model in a subset of our cohort. In a held-out test sample, we compared performance of our model to three clinical risk tools and one previously published model.

**Results:** Our 24-feature model achieved an area under the curve (AUC) of 0.71 (95% confidence interval [CI], 0.69-0.72), higher than all other tools (research-based AUC: 0.67 [95%CI, 0.66-0.69], clinical AUCs: 0.55 [95%CI, 0.54-0.56] to 0.61 [95%CI, 0.59-0.62]). Five features were novel including red blood cell indices and infection markers measured standardly upon admission. Additionally, we identified inflection points for several vital signs and labs where risk rose substantially. Most notably, patients with median intrapartum systolic blood pressure above 132mmHg had an 11% [interquartile range, 4%] median increase in relative risk for PPH.

**Conclusions:** We developed a novel approach for predicting PPH and identified clinical feature thresholds that can guide intrapartum monitoring for PPH risk. Our results suggest our model is an excellent candidate for prospective evaluation and could ultimately reduce PPH morbidity and mortality through early detection and prevention.

## BACKGROUND AND SIGNIFICANCE

Postpartum hemorrhage (PPH) remains a leading preventable cause of maternal morbidity and mortality in the United States and worldwide[1–4]. Recent trends in the United States suggest rates of PPH are rising, with reported increase in prevalence from 2.9% in 2010 to 3.2% in 2014, which constitutes a 13.0% increase [5,6]. Though the risk of mortality has remained stable[4,7,8], PPH still represents 11.2% of maternal deaths in the United States[9], highlighting the need for accurate risk prediction and prevention.

PPH is most commonly defined using blood loss and clinical signs of hemodynamic compromise, though specific criteria vary[10]. The American College of Obstetrics and Gynecology (ACOG) reVITALize program defines obstetric hemorrhage in patients with a cumulative estimated blood loss >=1000mL or blood loss associated with signs or symptoms of hypovolemia within 24 hours of delivery[11]. Others have proposed different definitions, such as the World Health Organization, which defines postpartum hemorrhage as a cumulative blood loss of >=500mL within 24 hours after birth[12]. Differences and inconsistencies in definitions complicate identification and subsequent prediction of PPH[13].

Nonetheless, stratification tools based on known risk factors are used to identify women at high risk of obstetric hemorrhage, promoting clinical awareness and prompting measures to mitigate risk[11,14]. The California Maternal Quality Care Collaborative (CMQCC)[15], Association of Women’s Health, Obstetric and Neonatal Nurses (AWOHNN)[16], and New York Safety Bundle for Obstetric Hemorrhage (NYSBOH)[17] are widely used in the US at admission and during labor. These guidelines stratify women into low-, medium- and high-risk groups based on characteristics such as low platelet count, multiple gestation, prior Cesarean delivery or uterine surgery, and prior history of PPH to determine the need for pre-transfusion testing[18,19].

Evaluations of these stratification tools have revealed that they have limited clinical utility[18,20]. Assessments of the CMQCC have shown a statistically significant difference in risk for PPH between women in low-, medium- and high-risk groups. However, this same tool has consistently classified more than 40% of PPH cases as low-risk because they had no risk factors upon admission[18,21,22]. The tool underestimates risk for various definitions of severe PPH, as well as PPH defined as >1000mL cumulative blood loss. An assessment of the CMQCC, AWHONN and NYSBOH toolkits for predicting severe PPH (blood transfusion of >=4 units of blood) in women undergoing Cesarean delivery reported better performance with only 4%-17% of cases misclassified as low-risk[20]. While these cases are among the most critical to detect, they account for a relatively small proportion of total PPH cases. Nonetheless, modified versions of the CMQCC provide guidance nationally for pre-transfusion testing[11].

To improve on the accuracy of PPH risk prediction, novel approaches to predict PPH have utilized large datasets to identify more specific risk factors for PPH. Venkatesh and colleagues found that machine learning and statistical models utilizing data available at time of labor admission accurately predict PPH[23]. However, these types of models have not been systemically compared with risk stratification tools currently being used as part of standard of care.

Previously, we have developed and validated an accurate digital phenotyping algorithm to ascertain PPH from comprehensive electronic medical record (EMR) data that not only incorporates cumulative blood loss, but also other important diagnostic and treatment-related features indicating PPH, such as use of uterotonics and hemorrhage related procedures[24]. The combination of machine learning methods and EMR data allows for the construction of predictive models based on population-scale analyses that involve more precisely defined outcomes and exposures. We hypothesized that EMR data from our large, diverse health system may provide for a richer feature set to construct more predictive models for prospectively identifying those at risk for PPH. Here, we evaluate patients prior to delivery to predict their risk of PPH using integrated clinical features from large-scale, high-dimensional clinical data derived from the Mount Sinai Health System (MSHS) EMR database. Furthermore, we compare the performance of our model to existing risk assessment tools.

## MATERIALS AND METHODS

We aimed to build a novel informatics-based tool to assess risk of PPH. Figure 1 shows an overview of our study design. In prior work, we constructed a delivery cohort and developed and validated a high-accuracy PPH digital phenotyping algorithm to identify PPH cases retrospectively[24]. For a brief summary of this work, see the Supplementary Methods.

**Figure 1.**
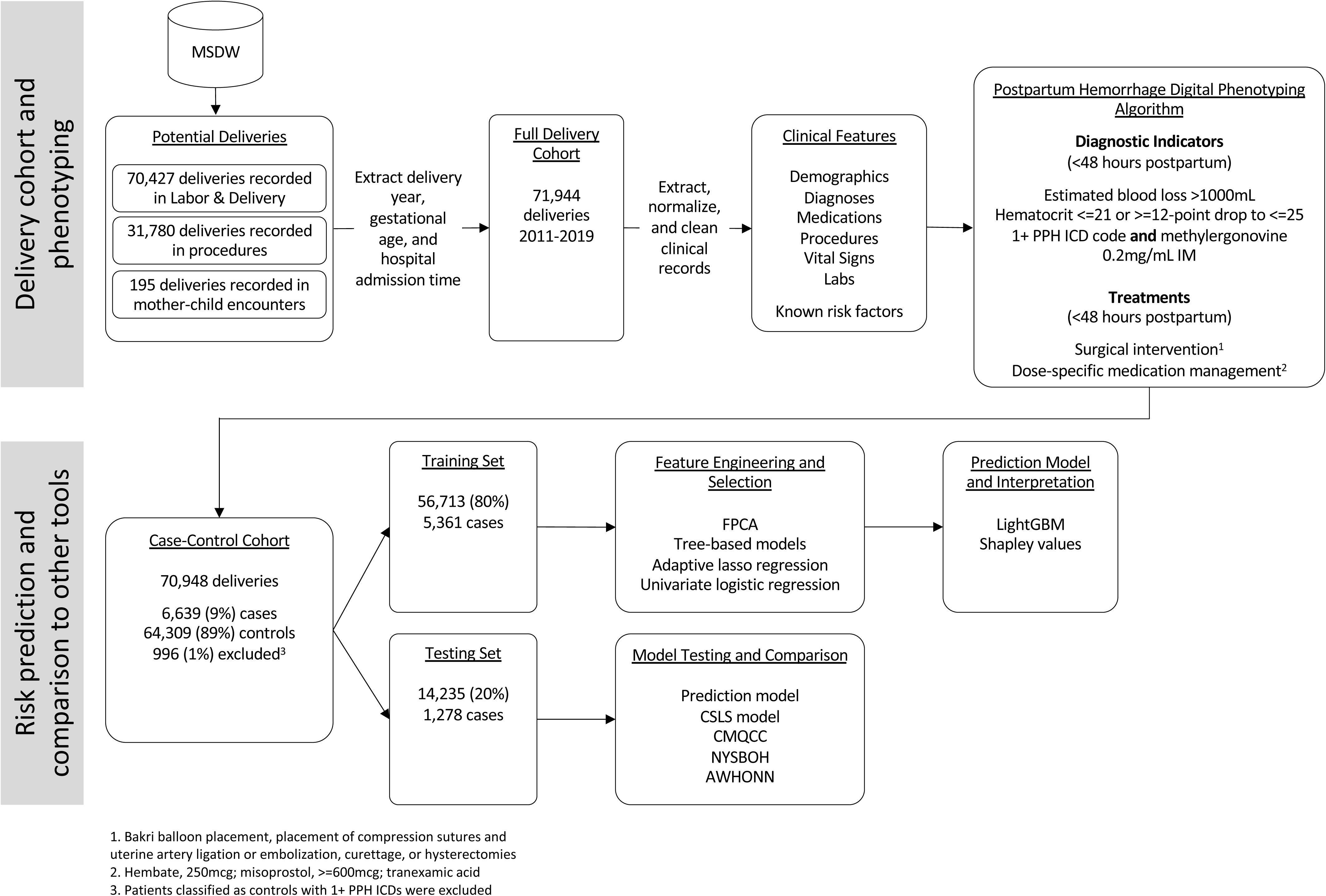
Overview of study design and model development

Here, we engineered thousands of potentially informative features from clinical information recorded prior to delivery for patients in our delivery cohort. Using PPH status determined by our digital phenotyping algorithm, we trained a model to estimate PPH risk prior to delivery, and then, on an independent test set, assessed performance of our model, a previously published model[23], and three widely used clinical risk toolkits[15–17]. Our approach differed from other tools in that we used data-driven approaches for feature engineering, selection, and model development, so we referred to it here as an ‘integrated machine learning’ (IML) model. We received approval from the Icahn School of Medicine at Mount Sinai Institutional Review Board (IRB-17-01245) to conduct this study.

### Experimental design

We used clinical information available prior to delivery time and up to eight months prior to pregnancy between January 1, 2011 and December 31, 2019. This time frame captures events occurring prior to and during pregnancy while limiting the variation in data availability across patients, which can often be substantial. By definition, PPH occurs after delivery, so we used delivery time as a natural cutoff for data inclusion and removed all data occurring at delivery time or later for all patients. Considering the particular value of clinical information immediately preceding delivery, including during labor (intrapartum), we split available medication, vital and lab data into two time periods: pre-hospital admission and post-admission (but still prior to delivery time). To allow for independent testing of our model and to facilitate comparison across other risk assessment tools, we divided our case-control cohort into training (80%) and testing (20%) sets. Mothers were randomly assigned to training or testing; all deliveries for a given mother were assigned to the same set.

### Feature engineering and selection

Current risk stratification efforts for PPH have relied primarily on known risk factors (e.g., prior Cesarean delivery)[20]. While these factors are significant predictors of PPH, in many cases, PPH occurs in women with no known risk factors[11,18,21]. Considering this, we included both known risk factors and thousands of potential predictors extracted from EMR to maximize our ability to detect any patterns of clinical information that increase risk for PPH. These predictors spanned the total set of disease categories, generic medications, procedures, lab values, and vital signs ever detected in our cohort and included multiple ways of measuring many factors (e.g., minimum and maximum pulse). Of note, we generated functional principal components (fPCs) to summarize multiple vital sign and lab measurements for each patient over time. We excluded any feature with fewer than five individuals with non-missing values since these features are unlikely to be informative. Among the remaining features, we used a combination of gradient boosting, adaptive lasso regression, and logistic regression to select a small subset of features for input into our risk model. Details for these steps were outlined in Supplementary Methods.

### Learning algorithm

Selected features were used to train gradient boosted decision tree models with 100-fold cross-validation employed within the training dataset with the python package LightGBM. Gradient boosting trains weak tree models sequentially, using prior errors to inform subsequent models, resulting in stronger performance. LightGBM is a particularly fast and accurate version of gradient boosting that builds trees leaf-wise (by branching just from the best leaf at each node) rather than level-wise. This approach is appropriate for EMR-derived features, given they are often sparse, have missing values, and can contain complex non-linear interactions. We used stratified k-fold splits to retain case-control rates across folds and balanced sample weights to boost the relative importance of cases in classification. We set iterations to continue only until there was no improvement in area under the receiver operating characteristic curve (AUROC) in validation. Within folds, the best model was selected based on the F1 score, which reflects the weighted average of precision and recall. We reported average sensitivity, specificity, positive predictive value (PPV), negative predictive value (NPV), AUROC, and F1 score across folds. The final model was estimated using the full training sample with the average number of best iterations from cross-validation and then applied without modification to the test dataset. We estimated 95% confidence intervals (95% CI) for test performance metrics via bootstrapping (1000 samples with replacement).

### Model interpretation and simplification

Knowing what information drives prediction is key for adoption by medical professionals. Here, we used Shapley values to estimate relative importance of each feature using the python package SHAP[25,26]. Shapley values measure the degree to which each feature pushes an individual’s predicted risk above or below the base value, where the base value represents the average predicted risk for the sample. Overall feature importance can be calculated by taking the mean across individual Shapley values for each feature. Shapley values can also be transformed to relative risk scores and plotted against patients’ values to show inflection points above or below which risk is substantially increased or decreased. Finally, we used Shapley values to simplify our model to the minimum necessary features for maximum performance to improve its clinical utility and ease of interpretation. See Supplementary Methods for more details.

### Comparison to other risk assessment tools

We extracted risk factors for three commonly used PPH risk assessments, the CMQCC, AWHONN, and NYSBOH[15–17]. As recommended, we assigned women to low-, medium-, or high-risk groups based on 16-17 largely overlapping binary criteria that evaluate medical history, obstetric complications, and current vital signs and labs[20]. Here, we used intrapartum versions when available as this timing better aligns with the timing of our model. We used both high and medium risk as cutoffs for computing sensitivity, specificity, PPV, NPV, and AUROC for each of the three toolkits.

We also implemented a previously published EMR-based risk prediction model which was based on the US Consortium for Safe Labor Study (CSLS), 2002-2008, as an additional comparison[23]. The authors used several statistical methods to combine 55 known risk factors. Here, we trained a gradient boosting machine (their highest performing method) using 54 of their 55 risk factors in our training sample and applied this model to our test sample for a direct comparison using the same performance metrics.

Details of risk factors for all tools were reported in Supplementary Methods. Significant differences between our model and all other risk assessment tools were assessed using a two-sided DeLong test for comparing AUROCs within the same sample[27].

### Comparison to an alternative phenotype

Since the PPH phenotype we used here is newly developed, we evaluated our model and all comparison risk assessment tools against a more commonly used phenotype, estimated blood loss (EBL) >=1000mL, as well. For these analyses, we followed the same procedures as described above, but using only patients with non-missing EBL values (N=63,348) and using EBL alone to define our outcome (deliveries with EBL >=1000mL were labeled cases; deliveries with EBL <1000mL were labeled controls).

## RESULTS

### Patient cohort demographics and clinical characteristics

Our final case-control cohort included 70,948 deliveries and PPH case prevalence was 9% (Figure 1). Cases and controls differed significantly in several demographic and clinical characteristics (Table 1). Additional summary statistics were reported previously[24].

**Table 1.**
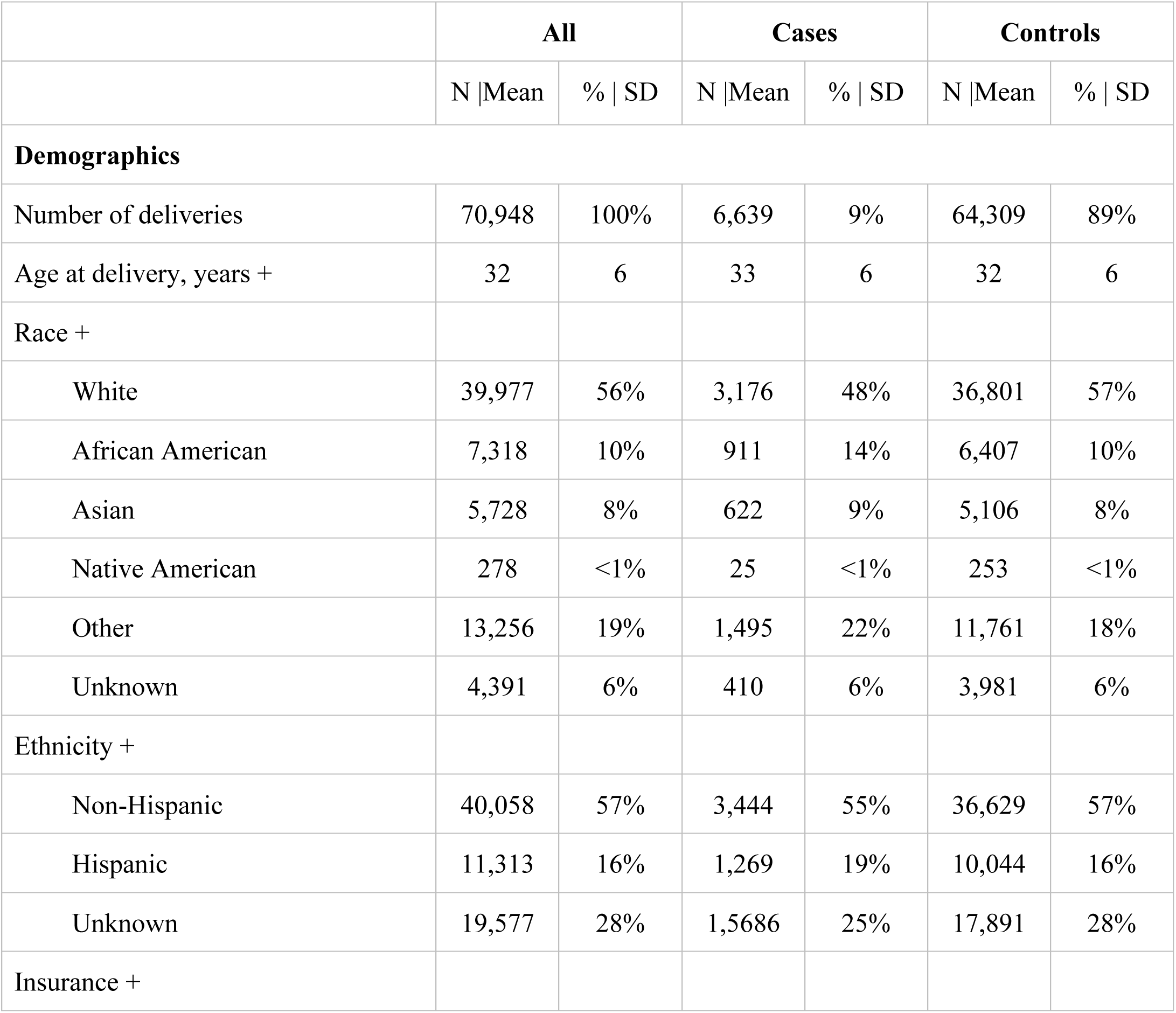
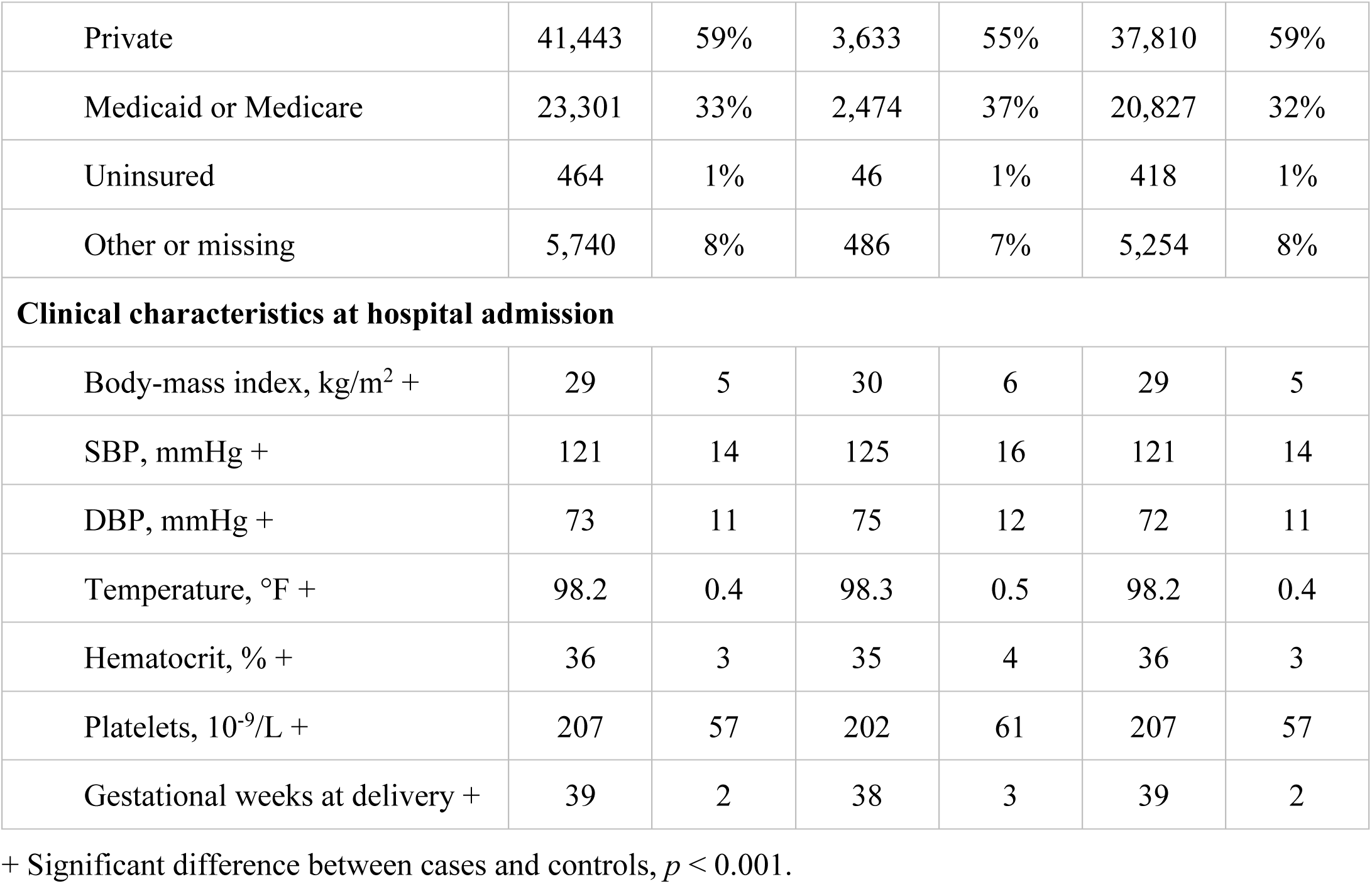
Demographics and clinical characteristics for the MSHS delivery cohort

### Selected features and model performance

We generated 5,327 features in our training sample, 3,982 of which had at least five non-missing values. Of these features, we selected 219 that showed significant association with PPH across two selection procedures (gradient boosting and either adaptive lasso or logistic regression). These 219 features represented 98 unique features (e.g., minimum and maximum pulse would be counted as two features, but only one unique feature).

All selected features were included in cross-validation training, but only 80 unique features (178 total features) received non-zero importance scores. Using these 80 features, our IML model achieved an AUROC of 0.73 (standard deviation [SD] = 0.03) in training and an AUROC of 0.72 (95% CI, 0.70-0.73) in the test set. Performance across subsets of features based on their importance scores suggested there was minimal additional value added beyond the top 29 features, representing 24 unique features. Using only these top 24 features, AUROC was 0.71 (95% CI, 0.69-0.72) in the test set. Additional performance metrics were listed in Table S1 (training) and Table 2 (testing).

**Table 2.** Vital signs and lab values show discrete increases in relative risk for PPH

| Feature during Hospital Admission | SHAP-based Cut Point | Increase in Relative Risk for PPH |  |  |
| --- | --- | --- | --- | --- |
|  |  | Quartile 1 | Median | Quartile 3 |
| Minimum SBP | 132 mmHg | 1.09 | 1.11 | 1.13 |
| Minimum DBP | 85 mmHg | 1.02 | 1.02 | 1.03 |
| Median pulse | 90 beats/minute | 1.03 | 1.04 | 1.05 |
| Minimum hematocrit | 30% | 1.02 | 1.05 | 1.07 |
| Minimum hemoglobin | 10.0 g/dl | 1.02 | 1.03 | 1.06 |
| Minimum platelets | $150 \times 10^9/L$ | 1.01 | 1.01 | 1.02 |

### Top 24 features highlight novel risk markers measured at admission and intrapartum

Among the top 24 unique features, 7 were lab results, 6 were diagnoses, 4 were vital signs, 4 were demographic variables, and 3 were medications. Considering the full list of 29 features, 19 features (66%) were ones used in current clinical practice or reported previously, including anemia, preeclampsia, and Cesarean delivery (Table S2). Five features (17%) were novel measures of risk factors previously reported. For example, we reported antepartum pulse as a top feature; pulse is a known risk factor, however, antepartum measures have not been previously considered. Finally, five features (17%) represent newly identified risk factors that may warrant further investigation including red blood cell count, mean corpuscular hemoglobin, red blood cell distribution width, absolute neutrophil count, and white blood cell count, many of which are measured in complete blood count panels. To illustrate how each unique feature contributed to individual predicted risk, we plotted patients’ Shapley values for the most important version of each of the top 24 unique features, excluding fPCs, colored by the feature value itself (Figure 2). Shapley values reflect the relative contribution of the feature to the patient’s predicted risk score; often high feature values had a larger impact on an individual’s risk score.

**Figure 2.**
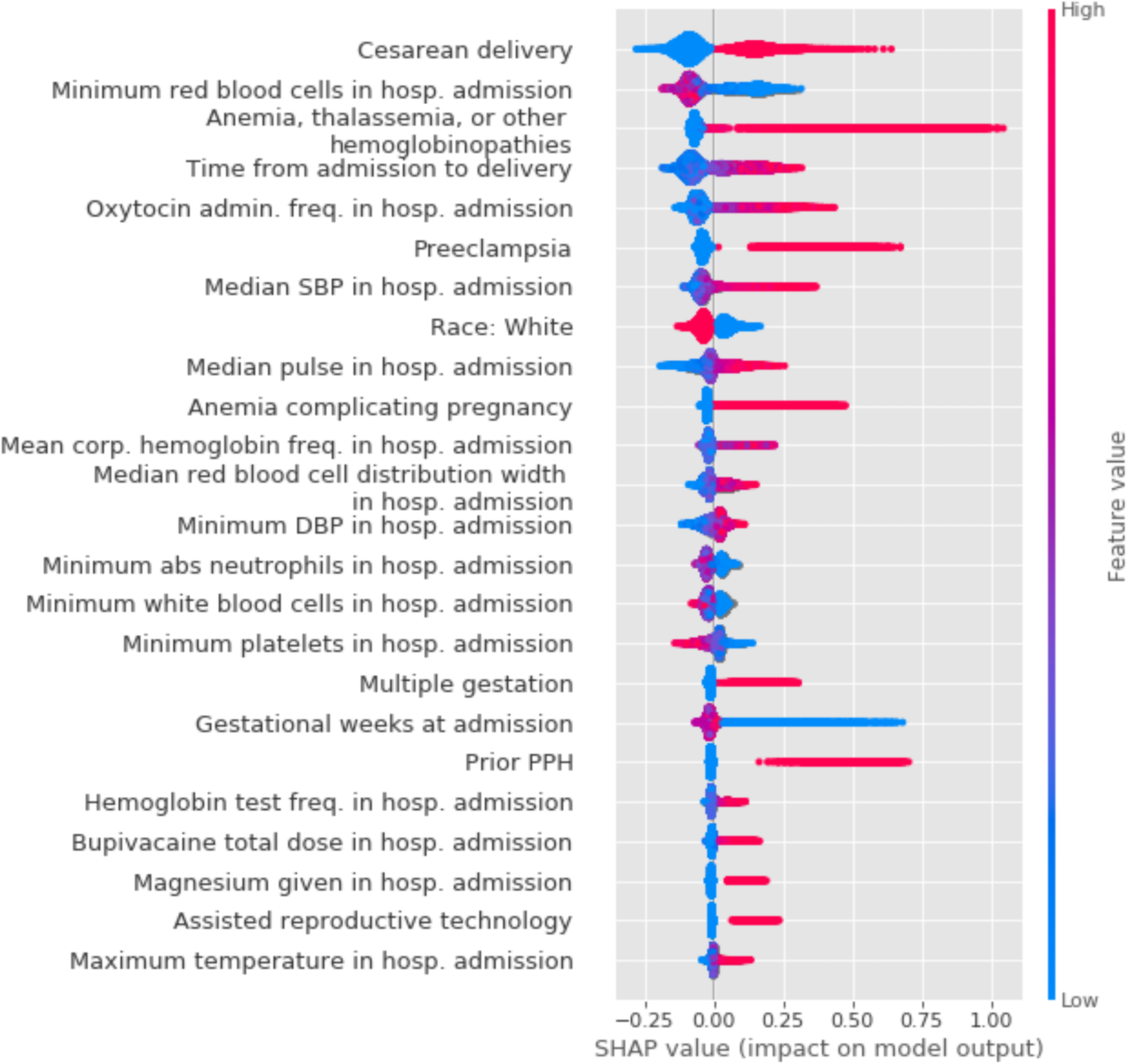
Shapley values for top 24 clinical features for prediction of PPH. Shapley values for the best version of the top 24 features were plotted in rank order of importance for patients in the training data. Positive Shapley values indicate an increase in risk for PPH and negative values indicate a protective effect. Points were colored based on the original feature values.

Blood pressure and pulse are measured frequently across the duration of hospital admission to monitor patient wellbeing. We calculated 3-hour moving averages and standard deviations for cases and controls during the 12 hours preceding delivery (Figure 3). Fixed effects models confirmed patients who developed PPH had consistently higher blood pressure and pulse in the hours prior to delivery (and hemorrhage) than patients who delivered without PPH (*p’*s <2 x 10-^16^, Table S3). Additionally, we found that relative risk (RR) for PPH increased, sometimes dramatically, when values for these key vital signs, as well as for the primary labs used for PPH risk assessment upon admission, passed certain inflection points (Table 2; Figure 4). As noted with the shaded areas in each plot, these points did not always align with the reference range for normal pregnancy values.

**Figure 3.**
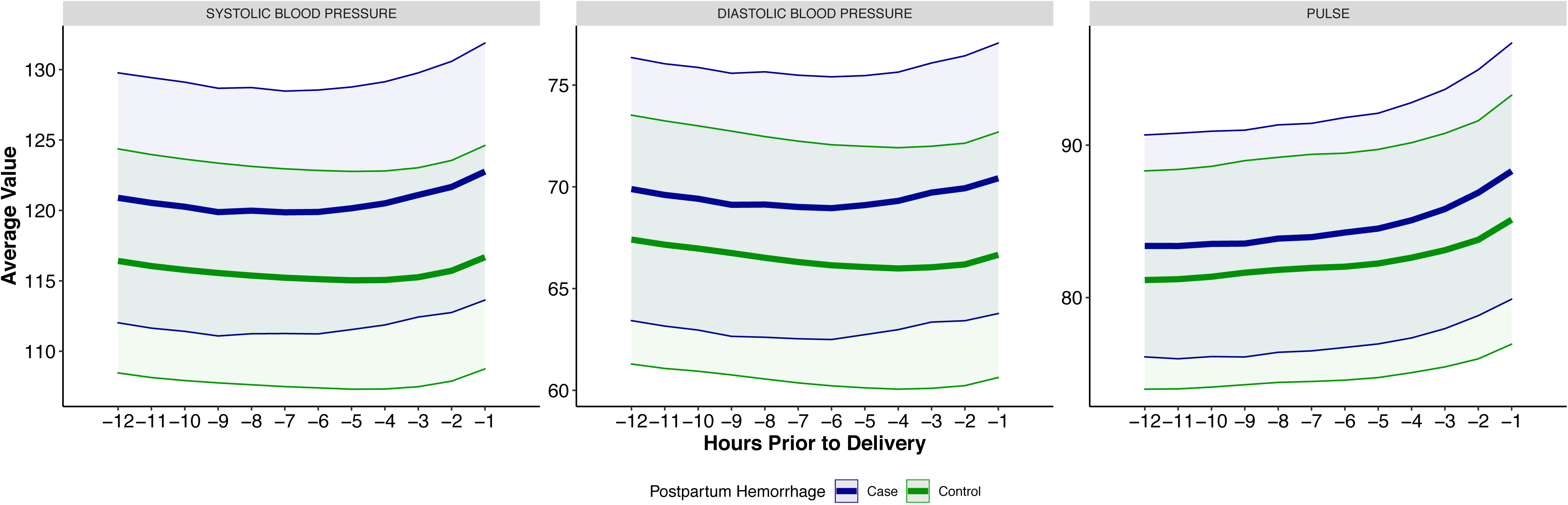
Dynamic changes of three vital signs consistently measured prior to delivery. Moving averages and standard deviations with 3-hour windows across the 12 hours prior to delivery were computed for cases and controls.

**Figure 4.**
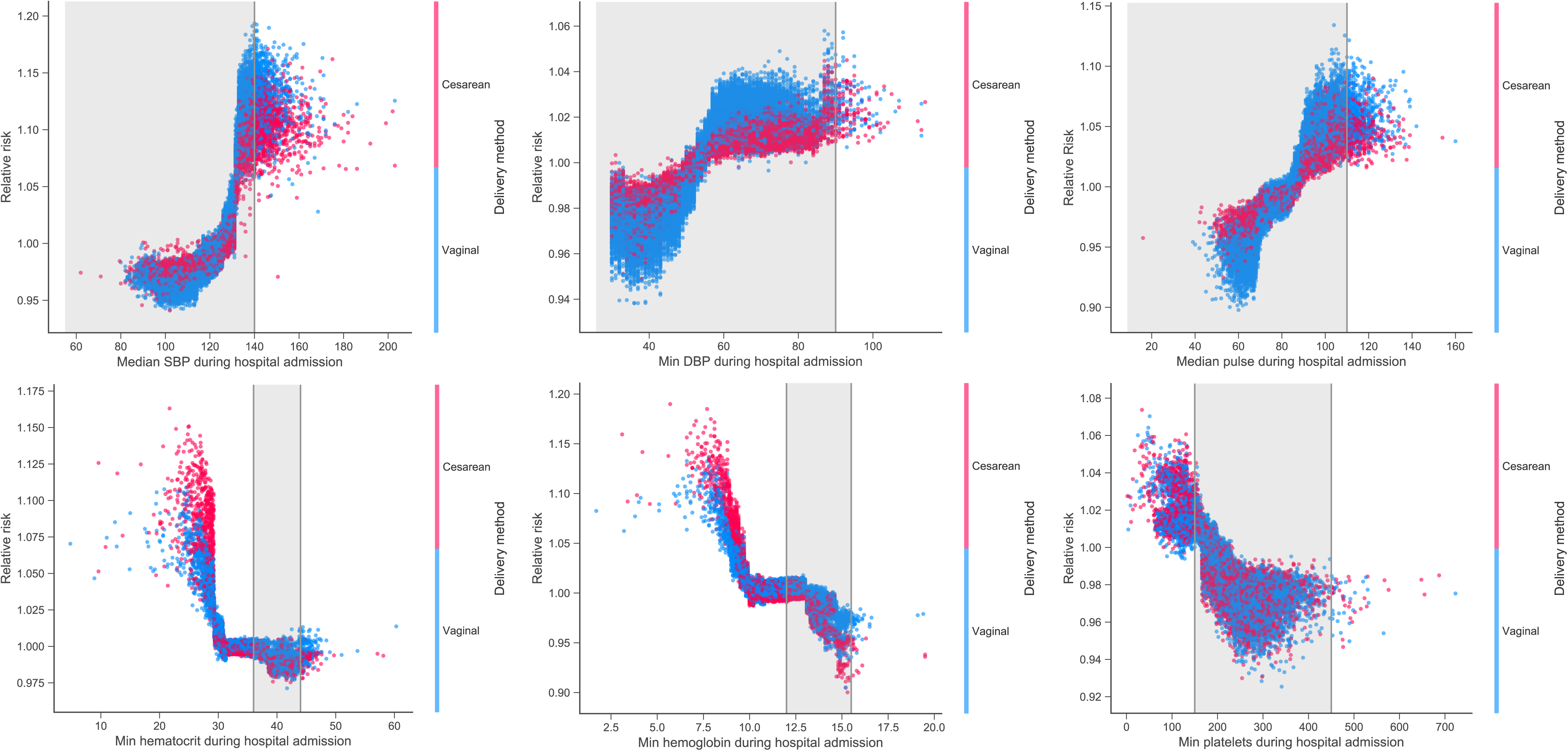
Relative risks for six vital signs and lab measurements stratified by type of delivery. Relative risk for PPH was plotted against feature values for patients in the training data for important vital signs and lab measures. Shaded gray area reflects the reference range for normal values in pregnancy. Points are colored by the delivery method (Cesarean or vaginal).

### Predictive risk model outperforms previously published and existing risk assessment tools

Our 24-feature IML model achieved a significantly higher AUROC than all other risk assessment tools we applied (Table 3, *p’*s <1.94 x 10^−7^ from two-sided DeLong test). The CSLS model achieved an AUROC of 0.67 (95% CI, 0.66-0.69) and the clinical risk assessment toolkits yielded AUROC values between 0.55 (95% CI, 0.54-0.56) and 0.61 (95% CI, 0.59-0.62) across toolkits and case classification thresholds (Table 3). Since the CSLS and our models had no clear risk category thresholds, we assigned risk category labels using deciles of predicted risk (top 10% = high-risk; 60-90% = medium-risk, <60% = low-risk) to compare precision across risk tools (see Table S4). Among the high-risk category, PPV was 28% for our model, 24% for the CSLS model, and 15-19% for the clinical toolkits (Figure 5).

**Table 3.** Performance matrix across risk assessment tools in our independent test set

| **Risk Assessment Tool** | **Sensitivity** | **Specificity** | **PPV** | **NPV** | **AUROC** |
| --- | --- | --- | --- | --- | --- |
| Integrated Machine Learning (all 80 vars) | 0.57<br>[0.54-0.60] | 0.73<br>[0.72-0.74] | 0.17<br>[0.16-0.18] | 0.95<br>[0.94-0.95] | 0.72<br>[0.70-0.73] |
| Integrated Machine Learning (top 24 vars) | 0.58<br>[0.55-0.61] | 0.71<br>[0.70-0.72] | 0.17<br>[0.15-0.18] | 0.95<br>[0.94-0.95] | 0.71<br>[0.69-0.72] |
| Consortium for Safe Labor Study | 0.56<br>[0.53-0.59] | 0.69<br>[0.68-0.70] | 0.15<br>[0.14-0.16] | 0.94<br>[0.94-0.95] | 0.67<br>[0.66-0.69] |
| Intrapartum CMQCC – high risk | 0.27<br>[0.24-0.30] | 0.88<br>[0.88-0.89] | 0.19<br>[0.17-0.20] | 0.92<br>[0.92-0.93] | 0.58<br>[0.56-0.59] |
| Intrapartum CMQCC –<br>medium risk | 0.63<br>[0.59-0.68] | 0.58<br>[0.57-0.59] | 0.13<br>[0.12-0.14] | 0.94<br>[0.94-0.95] | 0.61<br>[0.59-0.62] |
| Intrapartum NYSBOH –<br>high risk | 0.22<br>[0.20-0.25] | 0.87<br>[0.87-0.88] | 0.15<br>[0.13-0.16] | 0.92<br>[0.91-0.92] | 0.55<br>[0.54-0.56] |
| Intrapartum NYSBOH –<br>medium risk | 0.60<br>[0.56-0.65] | 0.61<br>[0.60-0.62] | 0.13<br>[0.12-0.14] | 0.94<br>[0.93-0.94] | 0.60<br>[0.59-0.62] |
| Admission AWHONN –<br>high risk | 0.43<br>[0.40-0.47] | 0.75<br>[0.74-0.76] | 0.15<br>[0.13-0.16] | 0.93<br>[0.93-0.94] | 0.59<br>[0.58-0.61] |
| Admission AWHONN –<br>medium risk | 0.89<br>[0.84-0.94] | 0.20<br>[0.20-0.21] | 0.10<br>[0.09-0.10] | 0.95<br>[0.94-0.96] | 0.55<br>[0.54-0.56] |

**Figure 5.**
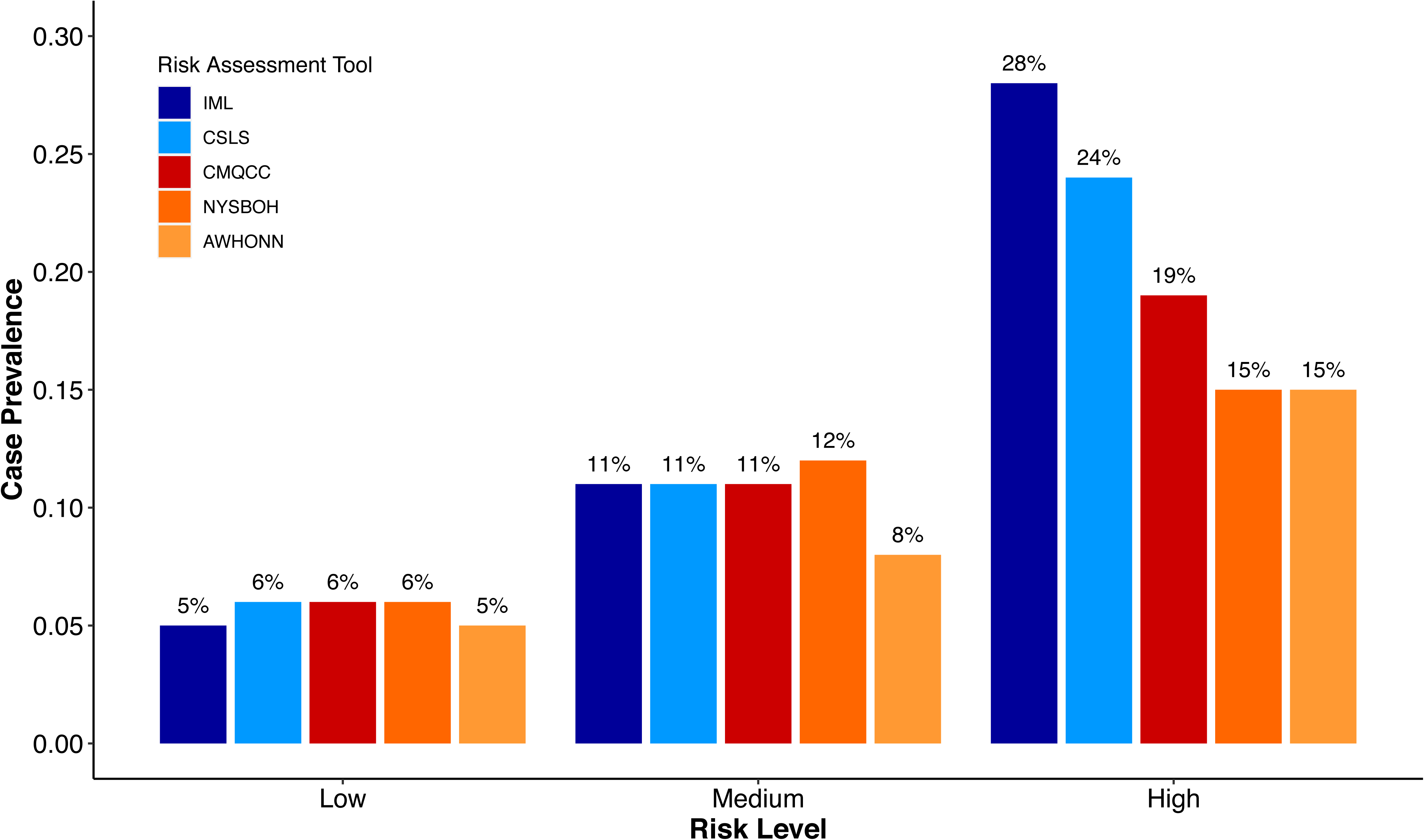
PPH prevalence among high-risk patients varies by risk assessment tools. Case prevalence within each risk category for each risk tool was calculated. Risk categories were assigned using deciles for CSLS and Sema4 models (high-risk = top 10%, medium-risk = 60-90%, low-risk = <60%).

Finally, we found a similar pattern of results using a more commonly implemented phenotype, EBL >=1000mL, rather than our digital phenotyping algorithm (Table S5). Case prevalence using EBL alone was 7% overall (N=4,182). Our IML model achieved an AUROC of 0.85 (95% CI, 0.84-0.87) using all 80 unique features, with no loss in AUROC when restricting the top 24 unique features (0.85, 95% CI 0.84-0.86); this was significantly higher than all other risk tools (*p’*s <2.2 x 10^−16^ from two-sided DeLong test). The CSLS model had an AUROC of 0.77 (95% CI, 0.75-0.78) and the clinical toolkits had AUROCs ranging from 0.55 (95% CI, 0.54-0.56) to 0.66 (95% CI, 0.64-0.57) depending on the toolkit and case classification threshold (Table S5).

## DISCUSSION

This study highlights a novel approach to predicting postpartum hemorrhaging utilizing a rich, diverse EMR dataset spanning nine years of deliveries. Our IML model used only 24 clinical variables – all passively collected through routine clinical care – to evaluate risk for PPH prior to delivery more accurately than three currently used risk assessment tools and a previously developed EMR-based model. When we used deciles of predicted risk to assign patients to risk categories, we found that PPH prevalence was 28% in our high-risk category, nearly twice as high as when risk was determined using clinical risk tools (15%-19%; Figure 5). Additionally, we identified inflection points for vital signs and labs where risk for PPH increased. These can help to guide risk management for PPH, while several novel risk factors may be additionally useful for monitoring risk during hospital admission. Finally, we showed that phenotype sensitivity can have a high impact on risk prediction research by comparing models using a highly accurate digital phenotype versus a less sensitive, blood loss-based one.

One significant advantage of our model is that it substantially outperformed currently used clinical tools with only 24 features, all of which are generally assessed prior to delivery. Our feature selection process was key to this success. All other risk assessment tools were based on expert opinion and clinical consensus, which does not always result in a set of features that maximize predictive accuracy. By assaying thousands of potential risk factors and using a suite of data-driven approaches to find the optimal set, we selected 24 known and novel risk factors that together delivered the highest performance of the tools we tested, including a model with more than twice as many features (CSLS, 55 features).

Our study also offered high-resolution data on vital signs and labs that are assessed and used for monitoring the peripartum period. In the obstetric population, there is a wide range of hemodynamic changes associated with blood loss[28,29]. Although hemodynamic changes encompass the reVITALize definition for PPH, there are no discrete definitions for these changes. Our model shows significant increasing trends for systolic blood pressure (SBP), diastolic blood pressure (DBP), and pulse in cases prior to delivery (Table S3). As depicted in Figure 3, SBP, DBP, and pulse begin rising at approximately 5 hours prior to delivery. There is significant variability in the relationship between blood loss and clinical signs, but these trends may provide useful insights for clinical care[29].

Additionally, we highlighted some notable transition points for important features such as platelets, hemoglobin, hematocrit, SBP, DBP, and pulse (Table 2; Figure 4). Notably, platelet count >150 appears to be protective of PPH. Recent data has shown that mild thrombocytopenia is an independent risk factor for PPH[30,31]. Nonetheless, our cohort provided robust data clearly showing an increasing risk of hemorrhage with decreasing platelet for both vaginal and Cesarean deliveries. Furthermore, we found that hematocrit and hemoglobin values reflected differential risks for PPH for Cesarean relative to vaginal deliveries. Risk for all patients increased with values lower than 30% or 10.0g/dl, respectively (Table 2), but the increase was steeper for Cesarean deliveries than vaginal deliveries (Figure 4). Finally, we found that risk increased at different thresholds for SBP, DBP, and pulse than what would be expected based on reference ranges for normal values (Table 2). For SBP, we observed an 11% median increase in risk for PPH when values were above 132 mmHg, despite a typical cutoff of <140 mmHg in pregnancy. Similarly, high pulse and DBP increased in risk at values within the normal range: 90 beats/minute and 85mmHg, respectively, while >110 beats/minute and >90mmHg are considered high in pregnancy. In general, our data suggests that intrapartum vital signs can capture risk for PPH and using PPH-specific risk guidelines may enhance monitoring relative to using reference ranges derived from the general population.

We also highlight several novel risk factors that are not currently monitored but are available from routine lab measurements and provide additional information for clinicians. We found red blood cells indices (red blood cell distribution width [RDW], red blood cell count, and mean corpuscular hemoglobin) were key risk factors for PPH in our model. RDW can be a marker for disease severity for underlying conditions such as diabetes, cardiovascular disease, and chronic kidney disease, as well[32,33]. These indices are likely associated with anemia, which impairs the physiologic response to blood loss and leads to worse prognosis. Further, infectious markers (white blood cell count and absolute neutrophil count) also had a high importance. These markers may indicate underlying inflammatory process, which also may alter or blunt the maternal response to blood loss leading to PPH. While these factors are already routinely assessed upon admission to Labor & Delivery, they are not currently used for monitoring risk. This may be a promising future direction for detecting individuals at risk, especially individuals presenting without other known risk factors.

Our final contribution was evaluating all risk prediction tools against a highly accurate PPH phenotype. Any assessment of the performance of a risk tool depends on how accurately the outcome it aims to predict is measured. Here, we compared performance of all risk tools for two definitions of PPH: 1) a digital phenotype that we developed[24], and 2) a phenotype based exclusively on a blood loss threshold for hemorrhage (>=1000mL) recommended by ACOG[11] and assessed by CSLS model[23]. We have previously compared these phenotypes to gold standard chart review labels and found the digital phenotype was significantly more accurate (AUROC of 0.85 vs. 0.67)[24]. A key driver of this discrepancy is that many patients with PPH do not pass this blood loss threshold (but can be identified using other measures of blood loss or receipts of treatment) and thus are misclassified as controls[13,24]. Here, using the same features and statistical methods for each tool, all risk models achieved higher performance for the EBL-based phenotype. Our model improved 14 points in AUROC (0.85 [95% CI, 0.84-0.86] vs. 0.71 [95% CI, 0.69-0.72]) by switching phenotypes. However, this high performance is not particularly meaningful given the low accuracy of the phenotype and underlines the importance of both developing high quality phenotypes and building risk assessment tools based on well-measured outcomes. Most prior work aimed at improving risk prediction for PPH has not benefited from using robust phenotypes and this remains a major barrier to achieving that goal.

Our study comes with several limitations: it is retrospective, relies on data from a single healthcare system, and does not utilize free-text notes. Our study may not reflect performance in a real-world clinical setting, and prospective validation of this work in diverse healthcare systems is critical to evaluate any potential clinical utility, although currently no such prognostic model exists for the general obstetric population[34]. Nonetheless, EMR in MSHS is the one of the largest and most comprehensive EMR systems, representing racial and ethnic diversity in New York City, as well as EMR implementation from various data sources. Finally, while our use of deidentified data can facilitate the application of our tools in research settings, systematic incorporation of data from clinical notes could bring valuable information particularly for patients missing key delivery details or without additional confirmation of PPH.

In summary, we utilized a large, robust, and diverse dataset to develop a novel risk prediction model for PPH, one of the leading causes of maternal mortality in the US. In comparison to existing risk assessment tools, including those currently used in clinical practice, we are able to achieve a higher AUROC with relatively few features using a highly accurate digital phenotype for PPH. We further identified inflection points for vital signs and lab values where risk for PPH begins to increase, which can be used as guidelines for monitoring risk intrapartum. The improvements in risk assessment afforded by our approach will require real-time, prospective evaluation in a hospital setting[35]. However, our results suggest using this model could facilitate early identification of PPH and allocation of appropriate resources[36], translating to reduced incidence, lower severity, and lower rates of maternal mortality.

## Data Availability

Data cannot be shared for ethical/privacy reasons.

## ACKNOWLEDGEMENTS

We would like to thank Mount Sinai Data Warehouse physician team for validating data accuracy and facilitating the chart review process. We would also thank the Sema4 IT team for infrastructural and computational support.

## DATA AVAILABILITY STATEMENT

Data cannot be shared for ethical/privacy reasons.

## Supplemental Methods

### Delivery cohort

We used deidentified EMR data from the MSHS, one of the largest and most comprehensive EMR systems in New York City. From the 9 million unique patients with any demographic or clinical data represented in this EMR, we derived a delivery cohort of 71,944 deliveries from 57,151 mothers occurring between January 1, 2011 and December 31, 2019. We identified deliveries using structured delivery summaries completed by the Labor & Delivery staff in the MSHS, procedure records, and encounter visit details. For all included deliveries, we extracted gestational weeks at delivery, delivery time, delivery method, and hospital admission time.

### Clinical data extraction, cleaning, and normalization

Demographics, lab results, vital signs, diagnoses, medications, and procedures occurring one year prior to pregnancy and up to one year after delivery were extracted for deliveries in our cohort. Data were standardized by mapping original names and values to common Unified Medical Language System (UMLS) frameworks to increase interoperability between healthcare systems and reduce dimensionality. All observations were cleaned and normalized within data type.

For patient demographics, we extracted mother’s age at delivery, race, ethnicity, and insurance. We selected the most common self-report when there were inconsistencies within a patient’s history of self-reported race or ethnicity. Lab test names and units were mapped to logical observation identifiers names and codes (LOINC). Values were cleaned (invalid results and text removed) and converted to numeric values or standardized to non-numeric ordinal scales depending on the test (e.g., ‘positive’ set to 1 and ‘negative’ set to 0). The earliest result was retained when there were duplicates (e.g., ‘preliminary’ and ‘final’ results from the same test with the same values). Vital signs, including weight, height, temperature, respirations, pulse, oxygen saturation, diastolic blood pressure (DBP), systolic blood pressure (SBP), and self-reported pain were standardized to common names and unit scales. Diagnoses from ICD-9-CM and ICD-10-CM were combined via mapping to single-level diagnosis categories using the Clinical Classifications Software[1]. When no timestamp was available for a diagnostic code (only a day), the timestamp was imputed to 11:59pm to avoid any future bias. We filtered medications to those administered (i.e., directly given to patients) and mapped the remaining medication names to RxNorm common ingredients regardless of brand or dose. Procedures were recorded through CompuRecord, an anesthesia information management program, using CPT-4 codes. When procedures included multiple timepoints (e.g., procedure start, anesthesia given, fluid given), only the earliest one was retained.

### Digital phenotype algorithm for postpartum hemorrhage

Deliveries followed by postpartum hemorrhage were identified using a previously developed, rule-based digital phenotype that was validated against gold standard labels from physician chart review (89% accuracy). The phenotype combines diagnostic and treatment information, which yielded higher accuracy than using blood loss alone to designate PPH status (65-67% accuracy, depending on the cutoff)[24]. Specifically, any of following criteria was sufficient for PPH classification, 1) cumulative blood loss was >1000mL, 2) hematocrit (a proxy measure for blood loss) was critically low (<=21%) or dropped substantially (<=12-point drop from admission baseline to <=25%) within 48 hours of delivery, 3) receipt of PPH-specific medications including carboprost tromethamine, misoprostol, or tranexamic acid within 48 hours of delivery, 4) one of 30 PPH ICD-9/10 diagnostic codes in addition to methylergonovine within 48 hours of delivery (a medication given less specifically for PPH), or 5) any surgical intervention for PPH including Bakri balloon placement, placement of compression sutures and uterine artery ligation or embolization, curettage, or hysterectomies within 48 hours of delivery. For additional details on phenotype development and validation, please see the original publication[24]. Significant differences between cases and controls on demographic and clinical characteristics were determined using chi-square tests for independence (proportional differences) and logistic regressions (continuous measurements).

### Feature engineering

#### Diagnoses, procedures and medications

For diagnoses, we set a patient’s value to the earliest week they received any ICD-9/10 in a given disease, defined using the approximately 280 clinical classification software (CCS) single-level diagnosis categories[2–4], where week one was eight months prior to pregnancy. For any given diagnosis category, if a patient did not receive a code in that category, their value was set to zero. The same approach was used for procedures (CPT-4 codes) and medications given prior to hospital admission for delivery (any medication brand or dose mapped to the generic drug). This strategy allowed us to retain initial timing information in addition to presence or absence of the feature.

For medications given during hospital admission for delivery, we quantified the frequency of administration (unique timestamps between admission and delivery times), as well as the total dose for the sixteen most commonly given drugs. We limited dosing variables to the most common for a number of reasons: 1) it was not available for every administration, 2) it required significant additional standardization across units, and 3) variation across patients is limited among medications given less commonly in a relatively short time window (admission to delivery).

#### Vital signs

For vital signs (excluding pain), we generated summary features for each of the two time periods (prior to and after admission) including minimum, median, and maximum values. We also generated 10 functional principal components using the R package fdapace for each vital sign within each time period using the maximum daily value for each patient when there was more than one. Time was standardized across individuals by taking the difference between the measurement day and the first day of the pregnancy. Functional principal component analyses (fPCA) construct eigenvectors that best represent the covariance matrix where each observation is a time series of values (for example, a patient’s temperature across pregnancy). Individual scores for each fPC were generated for all patients based on fPCAs derived exclusively using the training data. For pain, the maximum value across physical locations for each time period was retained. We did not impute missing values for any vital signs.

#### Lab values

Numeric lab values were handled similarly to vital signs. We extracted the same summary values, as well as frequency, and generated fPCs as described above. However, since there were hundreds of lab tests, we only derived fPCs for 15 of the most common lab tests and we did not split values by time period because there were too few values post-admission. We additionally generated baseline values (the value with the earliest timestamp) and maximum change values (the largest shift from baseline during the period) for labs occurring post-admission. For nominal lab values, the maximum ordinal value for each time period was extracted. Missing lab values were left missing.

#### Known risk factors

We manually curated 59 variables spanning demographics, obstetric characteristics, admission baseline vital signs, admission baseline hematocrit and platelet lab values, pregnancy complications, and general medical history that were included in current risk toolkits or used in previously published risk models for inclusion in our feature selection process. The demographic variables we used were age at admission, race (Black, Asian, White, Native American, Other), ethnicity (Hispanic or Latino, Not Hispanic or Latino), insurance (Medicaid or Medicare, private insurance, uninsured, other insurance or missing), alcohol history, and tobacco history. Obstetric characteristics included gestational weeks at admission, dummy variables for gestational weeks <32, between 32 and 37, and >37, delivery method (Cesarean or vaginal), labor trial, spontaneous labor, induced labor (as indicated by oxytocin administration records), parity, a dummy variable for parity >4, prior preterm birth, multiple gestation, and time from admission to delivery. For baseline admission vital signs and labs, we extracted the first available value after admission for SBP, DBP, temperature, height, weight, hematocrit, and platelets. We additionally had dummy variables for baseline hematocrit <30, baseline platelets <100,000, and baseline platelets <70,000. Body-mass index (BMI) at admission, as well as prior to pregnancy, were included, as well as dummy variables for admission BMI >35 and BMI >40. Weight prior to pregnancy was also included.

For pregnancy complications, obstetric history, and medical history, we used binary variables based on ICD codes and CPT-4 procedure codes. The following conditions were included: assisted reproductive technology, breech presentation, chorioamnionitis during hospital admission, early false labor (<37 weeks), eclampsia, fibroids, group B streptococcal infection, large for gestational age (antenatal diagnosis), preeclampsia (measured using ICDs as well as using a more comprehensive digital phenotype), severe preeclampsia, superimposed preeclampsia, placental abruption, placenta accreta, placenta previa, polyhydramnios, prolonged second stage of labor, premature rupture of membranes (PROM), small for gestational age (antenatal diagnosis), vaginal bleeding, prior miscarriage or abortion, prior PPH, prior stillbirth, anemia, asthma, coagulopathy, depression, diabetes, gastrointestinal disease, gestational diabetes, heart disease, chronic hypertension, renal disease, seizures, and thyroid disease.

### Feature selection

Among the thousands of features with five or more non-missing values, we used three strategies to select a subset of features for input into our risk model in order to avoid overfitting. First, we used the python package XGBoost to estimate feature importance within type (diagnoses, procedures, medications, vital signs, labs). To get a robust estimate of feature importance, we used the 75^th^ percentile importance score for each feature across 100 bootstrapped (60% randomly sampled per run) gradient boosted decision tree models. We also used adaptive lasso (implemented through the R package caret) and univariate logistic regressions to subset features based on association with PPH. For diagnoses, procedures, and medications, we used coefficients from best adaptive lasso regression in five-fold cross-validation (i.e., lowest mean error) run within each type since these features have no missing values. For vital signs and labs, we used coefficients from univariate logistic regressions for each feature among patients with non-missing values that included race, ethnicity, age, smoking history, alcohol history, insurance type (private, public, other), and gestational weeks at admission as covariates.

Our final feature selection step considered information across these analyses. Features with non-zero importance scores from gradient boosting models that also showed significant association with PPH in either lasso or logistic regression (depending on the feature type) were retained. For lasso, we considered an association p-value < 0.01 as significant and for logistic regression, we considered a Bonferroni-corrected p-value < 0.05 adjusting for the total number of vital and lab features to be significant.

### Model interpretation and simplification

We used Shapley values to estimate relative importance of each feature using the python package SHAP[5,6]. Shapley values decompose the deviation from expected risk for each individual into the contributions of each feature. For some patient Y, the Shapley value for feature X is estimated by comparing patient Y’s predicted risk to their predicted risk when feature X is missing. This is actually relatively complex to estimate since a feature’s contribution to a non-linear, interactive model (such as those based on decision trees) depends on other feature values, so predictions generated from all possible subsets of features must be considered. In practice, Shapley values can be calculated efficiently using TreeExplainer, which takes advantage of the model’s known tree path structure to reduce the number of perturbations that must be evaluated. Once the patient-feature matrix of Shapley values has been generated, overall feature importance can be calculated by taking the mean across individual Shapley values for each feature.

It can also be informative to show dependency plots, which illustrate the relationship between patients’ values for a particular feature of interest and the corresponding increase in predicted risk attributable to that value. This can reveal inflection points above or below which risk is substantially increased or decreased. Since the raw output of LightGBM for classification is in logit (e.g., here, the raw predicted risk is the predicted log odds of PPH), Shapley values for our model were also in logit. To transform them to probabilities, we used a sigmoid function ! to compute relative risk (RR) as follows

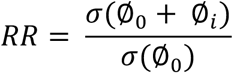

where ∅_0_ is the base Shapley value and ∅*_i_* is the Shapley value for a feature of interest, reflecting the degree to which that feature increased or decreased predicted risk from the base value for each patient[7].

Towards the goal of building an interpretable, stream-lined model for predicting risk, we tested the performance of our model using only the most important features. We tested our model using only the top feature and then sequentially adding the next important feature across the entire feature set to determine the minimal number of features needed for maximum AUROC. For each feature subset, we performed 10-fold cross-validation in our training data using the same parameters as our main LightGBM models and extracted the same performance metrics. This allowed us to quantify the marginal performance increase of each additional feature and find the inflection point at which adding more features yielded insufficiently small performance improvements.

### Risk factors used in other risk assessment tools

CMQCC, NYSBOH, and AWHONN risk assessment toolkits assign women to low-, medium-, or high-risk based on presence or absence of 16-17 variables[2–4]. The criteria for each toolkit were made available by Kawakita and colleagues and for CMQCC, we also included high-risk variables listed as intrapartum risk factors[5,6]. To implement these classifications here, we used any potential indication of the condition (ICD codes, procedures, medications, lab values, vital values) available prior to delivery.

For intrapartum assessment using the CMQCC, women with placenta previa, placenta accreta, active bleeding upon admission, platelets <100,000, a known coagulopathy, prolonged second stage of labor, prolonged oxytocin administration (>24 hours), magnesium sulfate during admission, or hematocrit <30 combined with any other risk factor (including those for medium risk) were considered high-risk. Women without any of those risk factors, but who have had a prior Cesarean delivery or other uterine surgery, multiple gestation, more than four previous vaginal births, chorioamnionitis upon admission, prior PPH, large uterine fibroids, fetal weight >4 kilograms, or BMI >35 were considered medium-risk. Women without any risk factors were low-risk. NYSBOH labels were very similar to CMQCC, but with the following exceptions: high-risk included platelets < 70,000 (rather than 100,000) and women with two or more medium risk factors. Additionally, hematocrit <30 plus another risk factor, prolonged oxytocin (>24 hours), prolonged second stage of labor, and magnesium sulfate were all medium-risk factors instead of high-risk. Finally, AWHONN had similar high-risk criteria, but several additional medium-risk criteria. Women with placenta previa, placenta accreta, active bleeding upon admission, platelets <100,000, a known coagulopathy, more than one prior PPH, hematocrit <30 combined with any other risk factor (including those for medium risk), or two or more medium risk factors were considered high-risk. Women without any of those risk factors, but who have had a prior Cesarean delivery or other uterine surgery, multiple gestation, more than four previous vaginal births, chorioamnionitis, prior PPH, large uterine fibroids, prior fetal demise, polyhydramnios, or induction of labor with oxytocin were considered medium-risk. Family history of PPH was also considered a medium risk factor, but could not be extracted from EMR data, so was excluded here.

We also used 55 features employed by Venkatesh and colleagues to predict PPH to train a model in our data using a very similar approach to theirs in order to compare their risk tool to ours[7]. Their features included age at admission, race, insurance, tobacco history, marital status, education status, recreational drug use history, baseline DBP, SBP, temperature, and weight, pre-pregnancy weight and BMI, gestational weeks at admission, multiple gestation, parity, labor trial, spontaneous labor, assisted reproductive technology, breech presentation, chorioamnionitis upon admission, early false labor (<37 weeks), eclampsia, group B streptococcal infection, large for gestational age (antenatal diagnosis), preeclampsia, severe preeclampsia, superimposed preeclampsia, placental abruption, placenta accreta, placenta previa, polyhydramnios, PROM, small for gestational age (antenatal diagnosis), vaginal bleeding, miscarriage or abortion, prior stillbirth, prior preterm birth, prior Cesarean delivery, anemia, asthma, coagulopathy, depression, diabetes, gastrointestinal disease, gestational diabetes, heart disease, chronic hypertension, renal disease, history of seizures, thyroid disease, magnesium sulfate during hospital admission, antenatal steroids, and fetal macrosomia (fetal weight >4 kilograms). We did not have education status on a sufficient percentage of individuals to use, so we excluded this variable.

## Supplementary Results

**Table S1.**
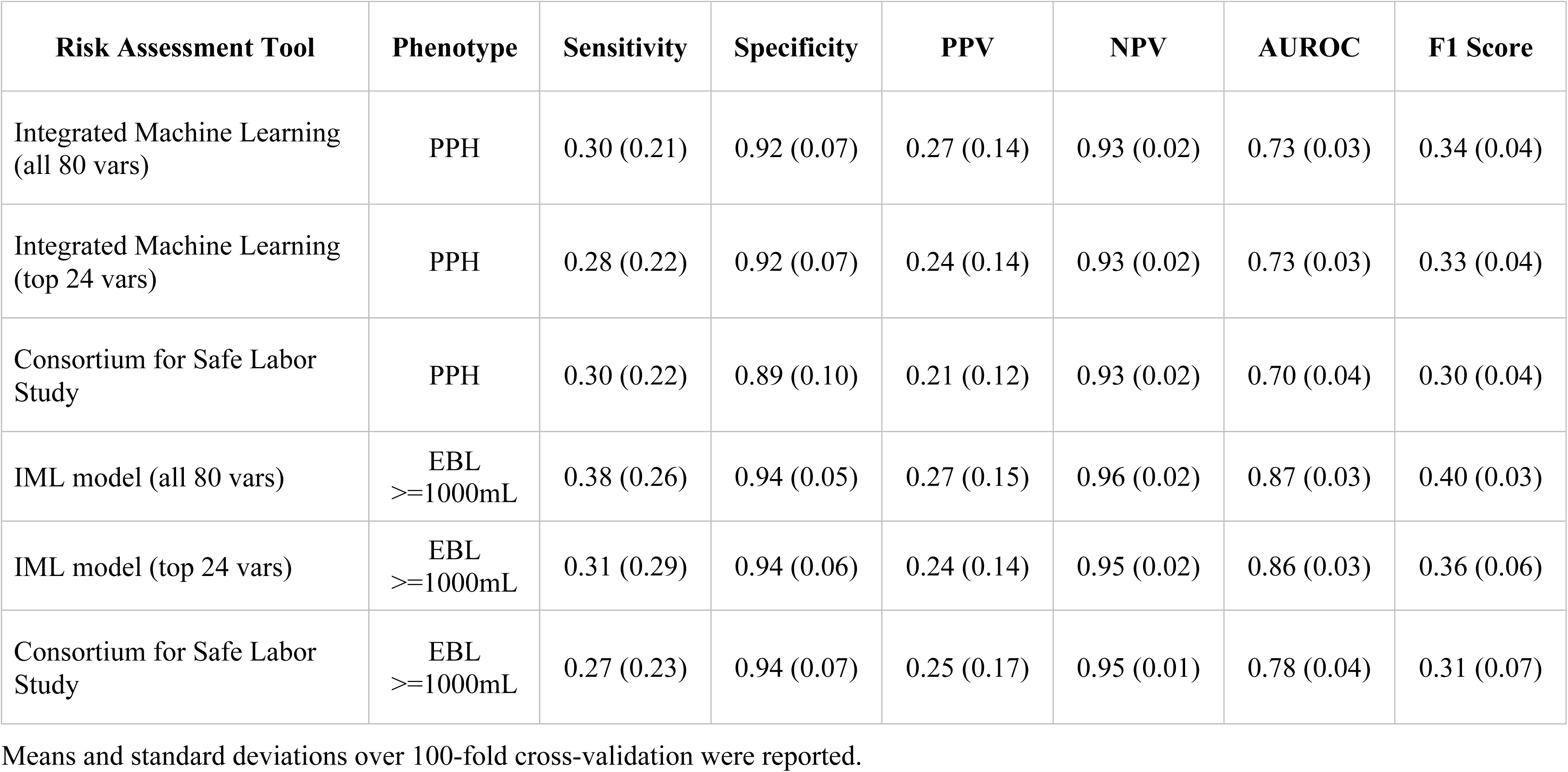
Training performance for machine learning risk assessment tools

| **Risk Assessment Tool** | **Phenotype** | **Sensitivity** | **Specificity** | **PPV** | **NPV** | **AUROC** | **F1 Score** |
| --- | --- | --- | --- | --- | --- | --- | --- |
| Integrated Machine Learning<br>(all 80 vars) | PPH | 0.30 (0.21) | 0.92 (0.07) | 0.27 (0.14) | 0.93 (0.02) | 0.73 (0.03) | 0.34 (0.04) |
| Integrated Machine Learning<br>(top 24 vars) | PPH | 0.28 (0.22) | 0.92 (0.07) | 0.24 (0.14) | 0.93 (0.02) | 0.73 (0.03) | 0.33 (0.04) |
| Consortium for Safe Labor<br>Study | PPH | 0.30 (0.22) | 0.89 (0.10) | 0.21 (0.12) | 0.93 (0.02) | 0.70 (0.04) | 0.30 (0.04) |
| IML model (all 80 vars) | EBL<br>≥1000mL | 0.38 (0.26) | 0.94 (0.05) | 0.27 (0.15) | 0.96 (0.02) | 0.87 (0.03) | 0.40 (0.03) |
| IML model (top 24 vars) | EBL<br>≥1000mL | 0.31 (0.29) | 0.94 (0.06) | 0.24 (0.14) | 0.95 (0.02) | 0.86 (0.03) | 0.36 (0.06) |
| Consortium for Safe Labor<br>Study | EBL<br>≥1000mL | 0.27 (0.23) | 0.94 (0.07) | 0.25 (0.17) | 0.95 (0.01) | 0.78 (0.04) | 0.31 (0.07) |

Means and standard deviations over 100-fold cross-validation were reported.

**Table S2.** Top clinical features include novel and known risk factors

| **Rank** | **Feature** | **SHAP Importance** | **Novelty** |
| --- | --- | --- | --- |
| 1 | Cesarean delivery | 0.119 | known |
| 2 | Minimum red blood cells in hosp. admission | 0.101 | novel |
| 3 | Anemia, thalassemia, or other hemoglobinopathies | 0.099 | known |
| 4 | Time from admission to delivery | 0.09 | known |
| 5 | Oxytocin admin. freq. in hosp. admission | 0.08 | alternate |
| 6 | Preeclampsia | 0.067 | known |
| 7 | Median SBP in hosp. admission | 0.061 | known |
| 8 | Race: White | 0.044 | known |
| 9 | Median pulse in hosp. admission | 0.043 | known |
| 10 | Anemia complicating pregnancy | 0.037 | known |
| 11 | Mean corp. hemoglobin freq. in hosp. admission | 0.033 | novel |
| 12 | Median red blood cell distribution width in hosp. admission | 0.032 | novel |
| 13 | Minimum SBP in hosp. admission | 0.026 | known |
| 14 | Minimum DBP in hosp. admission | 0.026 | known |
| 15 | Minimum absolute neutrophils in hosp. admission | 0.025 | novel |
| 16 | Minimum white blood cells in hosp. admission | 0.025 | novel |
| 17 | Minimum platelets in hosp. admission | 0.025 | known |
| 18 | Multiple gestation | 0.023 | known |
| 19 | Gestational weeks at admission | 0.022 | known |
| 20 | Temperature in hosp. admission [fPC #9] | 0.021 | alternate |
| 21 | Prior PPH | 0.019 | known |
| 22 | Maximum SBP in hosp. admission | 0.017 | known |
| 23 | Maximum pulse antepartum | 0.016 | alternate |
| 24 | Hemoglobin test freq. in hosp. admission | 0.015 | alternate |
| 25 | Bupivacaine total dose in hosp. admission | 0.014 | alternate |
| 26 | Minimum hemoglobin in hosp. admission | 0.014 | known |
| 27 | Maximum platelets in hosp. admission | 0.014 | known |
| 28 | Magnesium given in hosp. admission | 0.013 | known |
| 29 | Assisted reproductive technology | 0.013 | known |

Novelty was designated ‘known’ if it is currently used in clinical risk assessments or has been previously reported in the literature. ‘Alternate’ signifies that the risk factor is known, but the measure used here is novel. ‘Novel’ means the feature has not been previously reported as a risk factor for PPH.

**Table S3.** Linear fixed effects models for case differences in vital signs prior to delivery

|  | **Case Status**<br>**Estimate** | **Cluster**<br>**Standard**<br>**Error** | ***t*** | ***p*** |
| --- | --- | --- | --- | --- |
| Systolic blood pressure | -5.19 | 0.26 | -19.95 | $2 \times 10^{-16}$ |
| Diastolic blood pressure | -3.08 | 0.18 | -16.93 | $2 \times 10^{-16}$ |
| Pulse | -2.5 | 0.23 | -10.90 | $2 \times 10^{-16}$ |

Linear fixed effects models were run for each vital sign with case status and hour prior to delivery (set as a categorical variable for hours -13 to -1). Standard errors were clustered by delivery to account for serial correlation across time.

**Table S4.** Risk categories across risk assessment tools

| **Risk Assessment Tool** | **High-Risk** | **Medium-Risk** | **Low-Risk** |
| --- | --- | --- | --- |
| Integrated Machine Learning (all 80 vars) | 1,423 (10%) | 4,269 (30%) | 8,543 (60%) |
| Consortium for Safe Labor Study (CSLS) | 1,423 (10%) | 4,269 (30%) | 8,543 (60%) |
| Intrapartum CMQCC | 1,862 (13%) | 4,381 (31%) | 7,992 (56%) |
| Intrapartum NYSBOH | 1,935 (14%) | 3,937 (28%) | 8,363 (59%) |
| Admission AWHONN | 3,799 (27%) | 7,649 (54%) | 2,787 (20%) |

Number of patients and percent of test set were listed. For the CSLS and our models, high-risk was set to the top 10% of predicted risk, medium-risk was set to the following 30%, and low-risk was assigned to the remaining deliveries. Clinical risk tools (CMQCC, NYSBOH, and AWHONN) exclusively assign category labels (rather than a continuous probability) using presence of absence of risk factors.

**Table S5.** Test performance across risk assessment tools for EBL phenotype

| **Risk Assessment Tool** | **Sensitivity** | **Specificity** | **PPV** | **NPV** | **AUROC** |
| --- | --- | --- | --- | --- | --- |
| Integrated Machine Learning (all 80 vars) | 0.85<br>[0.83-0.87] | 0.73<br>[0.72-0.74] | 0.19<br>[0.17-0.20] | 0.99<br>[0.98-0.99] | 0.85<br>[0.84-0.87] |
| Integrated Machine Learning (top 24 vars) | 0.85<br>[0.83-0.88] | 0.72<br>[0.71-0.73] | 0.18<br>[0.17-0.19] | 0.99<br>[0.98-0.99] | 0.85<br>[0.84-0.86] |
| Consortium for Safe Labor Study | 0.70<br>[0.67-0.72] | 0.71<br>[0.70-0.71] | 0.15<br>[0.13-0.16] | 0.97<br>[0.97-0.97] | 0.77<br>[0.75-0.78] |
| Intrapartum CMQCC – high risk | 0.26<br>[0.22-0.30] | 0.88<br>[0.87-0.88] | 0.13<br>[0.11-0.15] | 0.94<br>[0.94-0.95] | 0.57<br>[0.55-0.58] |
| Intrapartum CMQCC – medium risk | 0.75<br>[0.69-0.80] | 0.57<br>[0.56-0.58] | 0.11<br>[0.10-0.12] | 0.97<br>[0.96-0.97] | 0.66<br>[0.64-0.67] |
| Intrapartum NYSBOH – high risk | 0.29<br>[0.26-0.33] | 0.87<br>[0.86-0.88] | 0.14<br>[0.12-0.15] | 0.94<br>[0.94-0.95] | 0.58<br>[0.57-0.60] |
| Intrapartum NYSBOH – medium risk | 0.72<br>[0.66-0.78] | 0.59<br>[0.59-0.60] | 0.11<br>[0.10-0.12] | 0.97<br>[0.96-0.97] | 0.66<br>[0.64-0.67] |
| Admission AWHONN – high risk | 0.45<br>[0.41-0.50] | 0.74<br>[0.73-0.75] | 0.11<br>[0.10-0.12] | 0.95<br>[0.95-0.95] | 0.60<br>[0.58-0.61] |
| Admission AWHONN – medium risk | 0.91<br>[0.85-0.97] | 0.18<br>[0.18-0.19] | 0.07<br>[0.07-0.08] | 0.97<br>[0.96-0.97] | 0.55<br>[0.54-0.56] |

Test performance and bootstrapped 95% confidence intervals were reported.

